# Validating saliva as a biological sample for cost-effective, rapid and routine screening for SARS-CoV-2

**DOI:** 10.1101/2022.02.07.22269889

**Authors:** B.R. Ansil, Carolin Elizabeth George, Sindhulina Chandrasingh, Ashwin Viswanathan, Mukund Thattai, Padinjat Raghu, Santhosha Devadiga, Arun Geetha Harikumar, Pulleri Kandi Harsha, Indu Nair, Uma Ramakrishnan, Satyajit Mayor

## Abstract

**Purpose:** Compared to nasopharyngeal/oropharyngeal swabs, non-invasive saliva samples have enormous potential for scalability and routine population screening of SARS-CoV-2. In this study, we are investigating the efficacy of saliva samples relative to nasopharyngeal/oropharyngeal swabs for use as a direct source for the RT-PCR based SARS-CoV-2 detection.

**Methods:** Paired nasopharyngeal/oropharyngeal swabs and saliva samples were collected from suspected positive SARS-CoV-2 patients and tested using RT-PCR. Generalised linear models were used to investigate factors that explain result agreement. Further, we used simulations to evaluate the effectiveness of saliva-based screening in restricting the spread of infection in a large campus such as an educational institution.

**Results:** We observed 75.4% overall result agreement. Prospective positive samples stored for three or more days showed a drastic reduction in the probability of result agreement. We observed 83% result agreement and 74.5% test sensitivity in samples processed and tested within two days of collection. Our simulations suggest that a test with 75% sensitivity, but high daily capacity can be very effective in limiting the size of infection clusters in a workspace. Guided by these results, we successfully implemented a saliva-based screening in the Bangalore Life Sciences Cluster (BLiSC) campus.

**Conclusion:** These results suggest that saliva may be a viable sample source for SARS-CoV-2 surveillance if samples are processed immediately. We strongly recommend the implementation of saliva-based screening strategies for large workplaces and in schools, as well as for population-level screening and routine surveillance as we learn to live with the SARS-CoV-2 virus.

## Introduction

The COVID-19 pandemic spread rapidly in India, infecting more than 30 million people in two years [1]. Given this magnitude and speed, COVID-19 presents various diagnostic challenges, in the context of massive population density and limited diagnostic and health infrastructure capabilities. Viral diagnosis has progressed tremendously, and of the various modalities for SARS-CoV-2 diagnosis, the most reliable test is the reverse transcription-polymerase chain reaction (RT-PCR) on Nasopharyngeal/Oropharyngeal swabs collected in Viral Transport Medium (N/OPS-VTM). This demands skilled technical staff, involves procedural complexities such as viral inactivation and RNA extraction. Besides, the sample collection protocol causes significant discomfort to the patient [2] and demands strict protocols for prevention of infection to healthcare workers. These procedural complexities of the test are associated with increased cost and turnaround time.

The SalivaDirect is an alternative RNA extraction free, cost-effective RT-PCR-based protocol with a short turnaround time and less dependence on the supply chain [3]. Using saliva as a source sample has several advantages: 1) samples can be collected by patients without the help of trained personnel, 2) stringent personal protective equipment (PPE) are not required, 3) non-invasive routine testing is possible 4) dispensing with swabs or VTM adds flexibility, and 5) no requirement for RNA extraction reduces the cost and hence widens its applicability. Evidence related to using saliva for SARS-CoV-2 testing is evolving, and recent studies have reported promising results [4, 5]. Unfortunately, saliva-based tests in India are explored by very few studies [5, 6].

In this study, we assessed the performance of saliva relative to N/OPS-VTM samples for use as a direct source (without RNA extraction) for the RT-PCR based SARS-CoV-2 detection.

We also investigated correlates for discordance between N/OPS-VTM and saliva sample pairs. Further, we used simulations that incorporate concordance to evaluate the effectiveness of SalivaDirect in restricting the infection spread in a large campus such as an educational institution. Finally, we present a case study on the implementation of such a strategy in an educational institution. Through this study, we provide evidence for a low cost, easy, fast, and accurate test that has a considerable advantage in a country like India, especially in learning to ‘live with the virus ‘.

## Materials and Methods

### Ethical statement

The study was approved by the Institutional Review Board of Bangalore Baptist Hospital (BBB/IRB/2020/010); Institutional Human Ethics Committee and Institutional Biosafety Committee, National Centre for Biological Sciences (NCBS/IEC-22/01, NCBS/IEC-26/, NCBS:34IBSC/UR1).

### Sample collection, processing and testing for SARS-CoV-2

Samples were obtained from patients of Bangalore Baptist Hospital between December 2020 and May 2021. From each individual, a N/OPS-VTM and saliva (5 ml) were collected. Each paired sample was given a unique biorepository number and transported to the COVID-19 testing laboratory at Institute for Stem Cell Science and Regenerative Medicine (inStem). Upon arrival, samples were stored in a 4°C refrigerator in the biosafety laboratory. Storage time before processing the samples varied from 0 to 15 days with a mean of 4 days. In case of storage beyond two days, the samples were moved to a -20°C freezer.

N/OPS-VTM samples were processed following the Indian Council of Medical Research (ICMR) approved protocol. RNA was extracted using a magnetic bead-based automated Viral RNA Extraction protocol (Beckman Coulter Life Sciences). Saliva were processed following SalivaDirect protocol [3] and tested for RdRP, E and N genes of SARS-CoV-2 and human RNase P gene using NeoDx-CoviDx™ mPlex-4R SARS-CoV-2 RT-PCR Detection kits. Test results were evaluated based on manufacturer ‘s guidelines. Detailed sample collection and testing strategies are provided in the Supplementary material.

### Analysis

We determined (a) the number of individuals positive for SARS-CoV-2 in both N/OPS-VTM and saliva, (b) those positive only in N/OPS-VTM, (c) those positive only in saliva, and (d) those negative in both N/OPS-VTM and saliva. From this data, we computed the test sensitivity on each sample type. Since nasopharyngeal swab sampling has been shown to produce false negatives by RT-PCR [7], sensitivities for the saliva and the N/OPS-VTM are defined here respectively as (a+c)/(a+b+c) and (a+b)/(a+b+c), considering any individual with a positive result on one or the other sample as true positive [4].

We compared positive outcomes from the saliva results with true positives and used a Generalized Linear Model (GLM) to understand the factors that explain result agreement (or disagreement). We modelled result agreement/disagreement (success/failure) as a function of the sex of the patient, age, severity of the symptoms (asymptomatic and symptomatic), and storage duration (number of days between collection and testing; grouped into two bins: 0-2 days and 3-15 days), assuming a binomial error distribution. This analysis did not include individuals showed inconclusive results. We also assessed the Ct-value distribution of all test genes for concordant samples and compared using Wilcoxon test [8].

### Modelling infection spread on a network

We explored the effectiveness of campus-wide saliva-based screening using a Monte Carlo simulation of infection transmission in a network of 1600 individuals. In the simulation, a subset of individuals are tested each day, and test results are reported after some delay. Individuals who test positive are isolated, and their contacts are subsequently tested and isolated if positive. We start with a single positive case and run the simulation until there are no infected individuals remaining. The model is stochastic, so each run of the simulation produces a different result. The total number of infections at the end of the simulation defines the size of the cluster. An important goal of mitigation is to limit the size of a cluster seeded by a single infection. The model used for simulation is described in the Supplementary material.

## Results

### SARS-CoV-2 detection in paired N/OPS-VTM and saliva samples

We observed 75.4% overall result agreement between N/OPS-VTM and saliva sample pairs; 30.3% positive agreement and 45.1% negative agreement (Table 1). The inconclusive results were 1.1% and 5.7% for N/OPS-VTM and saliva, respectively. Interestingly, 3.4% of saliva samples were positive when the corresponding N/OPS-VTM were negative. The sensitivity of saliva and N/OPS-VTM were 70.2% and 92.9%, respectively. In the GLM with four variables, duration of storage was independently associated with result agreement (Supplementary Table 1). Among N/OPS-VTM positive or saliva positive patients, we observed a clear drop in result agreement (Fig 1) from 80.8% (SE 70.7-88) to 55.5% (SE 45.7-64.9) when samples were stored for more than two days (p=0.025). Concordance (83%) and the sensitivity (74.5%) of the saliva test improved when we considered only the samples tested within two days of collection (Table 2). This also resulted in a significant improvement (decrease) in inconclusive test results (1.8% in saliva and 0.9% in N/OPS-VTM). We also found that viral loads were statistically indistinguishable in positive sample pairs for all three viral genes (Wilcoxon p > 0.05, Fig 2) among positive concordant samples.

**Table 1:** A matrix showing the alignment of results from the two methods - N/OPS-VTM and Saliva - for all samples (N = 175). The asterisk indicate result agreements.

| N/OPS-VTM (N=175) |  |  |  |  |  |
| --- | --- | --- | --- | --- | --- |
| Saliva<br>(N=175) |  | Positive | Negative | Inconclusive | Total |
|  | Positive | 30.3% (53)* | 3.4% (6) | 0.6% (1) | 34.3% (60) |
|  | Negative | 14.3% (25) | 45.1% (79)* | 0.6% (1) | 60.0% (105) |
|  | Inconclusive | 5.7% (10) | 0.0% (0) | 0.0% (0) | 5.7% (10) |
|  | Total | 50.3% (88) | 48.6% (85) | 1.1% (2) | **175** |

**Table 2:** A matrix showing the alignment of results from the two methods - N/OPS-VTM and Saliva - for samples tested within two days of collection (N = 112). The asterisk indicate result agreements.

| N/OPS-VTM (N=112) |  |  |  |  |  |
| --- | --- | --- | --- | --- | --- |
| Saliva<br>(N=112) |  | Positive | Negative | Inconclusive | Total |
|  | Positive | 27.7% (31)* | 3.6% (4) | 0.9% (1) | 32.1% (36) |
|  | Negative | 10.7% (12) | 55.4% (62)* | 0.9% (1) | 67.0% (75) |
|  | Inconclusive | 0.9% (1) | 0.0% (0) | 0.0% (0) | 0.9% (1) |
|  | Total | 39.3% (44) | 59.0% (66) | 1.8% (2) | **112** |

**Fig 1:**
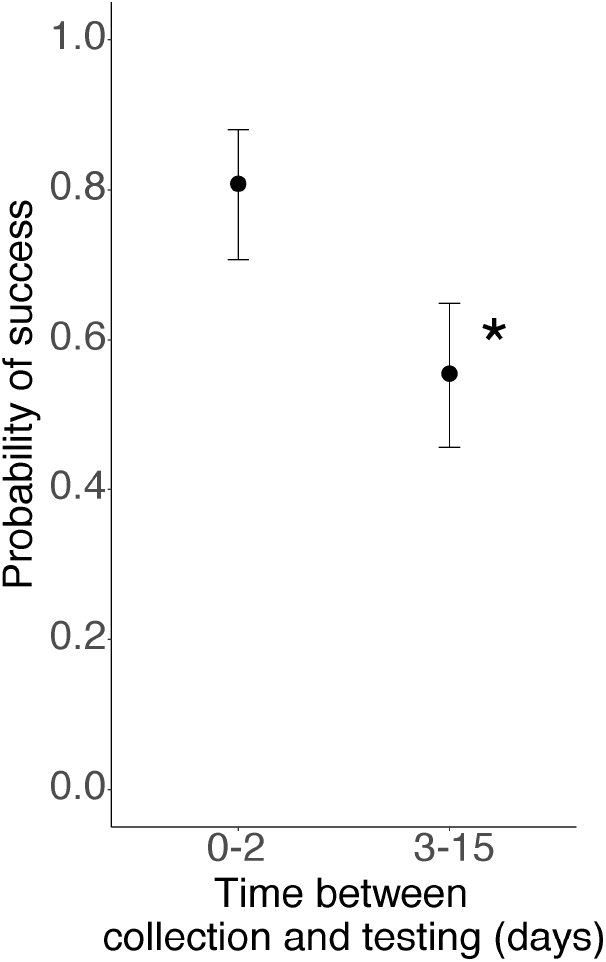
Probability of result agreement in positive samples (0-2 days: N = 49; 3-15 days: N = 46). Samples stored for more than two days showed high result disagreement between paired samples. Error bars are standard errors and the asterisk denotes a significant difference.

**Fig 2:**
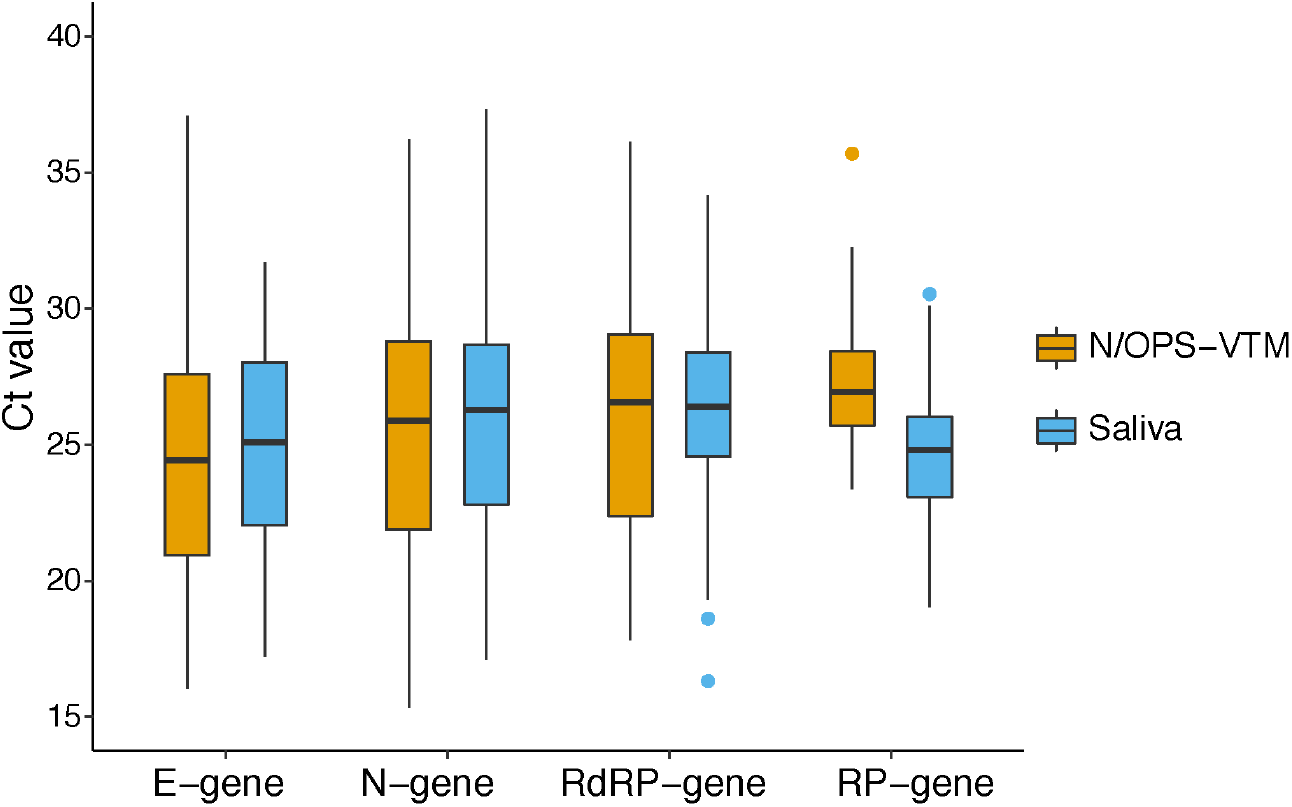
Boxplot showing Ct value distribution for three SARS-CoV-2 genes and human RNAse P gene. There is no significant difference in mean Ct values for viral genes between N/OPS-VTM and saliva samples.

### Simulations reveal that saliva-based screening can limit infection spread

We track the probability that a single starting infection leads to a large cluster (of size 25 or more) as the testing parameters are varied (Fig 3). This model is not meant to replicate the transmission dynamics of an actual workforce; rather, it is a proof of principle to identify key factors that influence the success of screening. We focus on three key factors of the testing protocol: (a) sensitivity, (b) daily testing capacity, (c) delay in reporting results. In our simulations, in the absence of testing and isolation, the probability of a large cluster is 20% under the assumed parameter values. However, by using a test with 75% sensitivity, at 200 tests per day with a one-day delay for results, we decrease the probability of a large cluster ten-fold, to about 2%. Moreover, this protocol works better than a test with 100% sensitivity, but with half the capacity or twice the delay.

**Fig 3:**
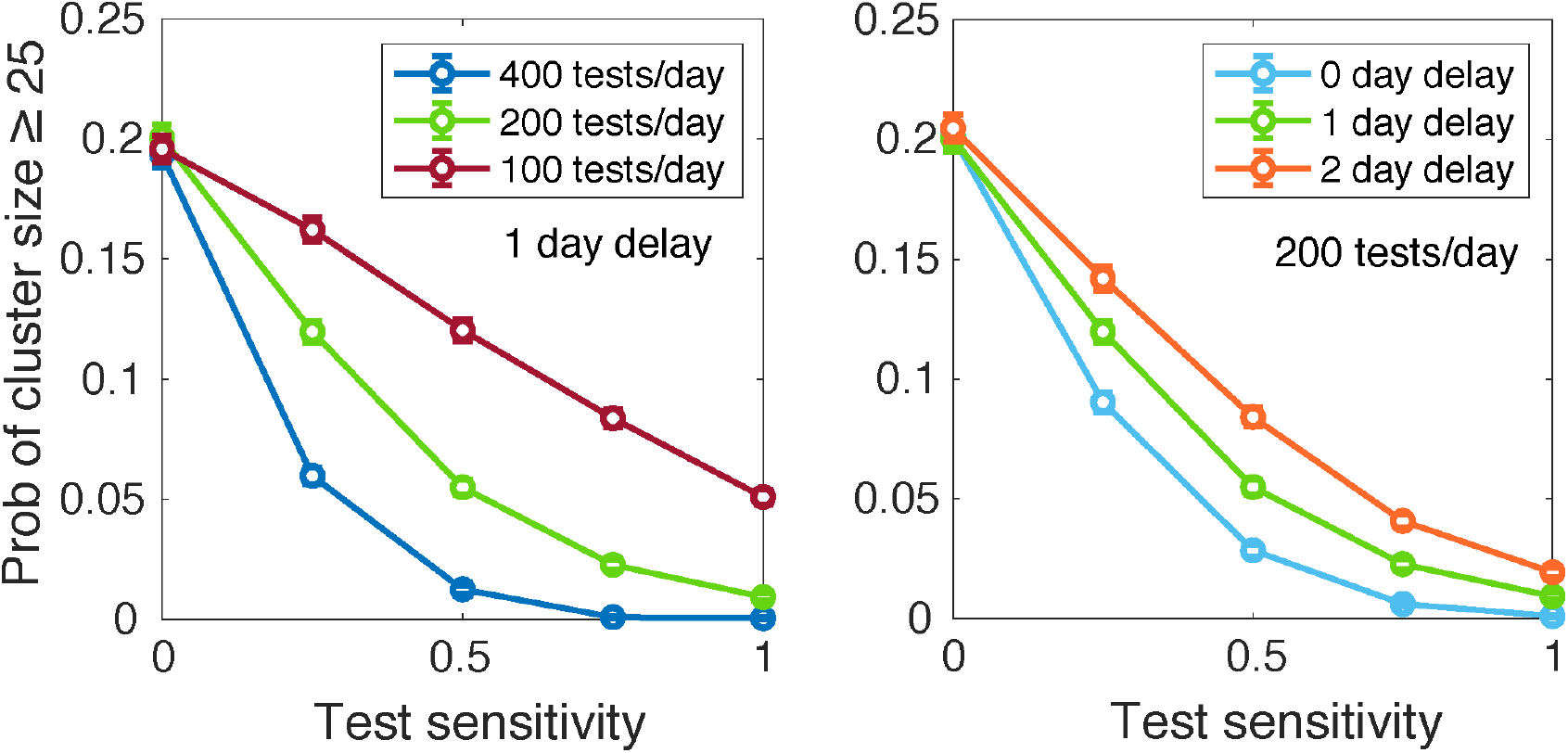
Results of a Monte Carlo simulation of infection transmission on a network of 1600 individuals. We track the probability that a single starting infection seeds a cluster of 25 or more infections. We compare a baseline protocol that has a capacity of 200 tests per day and a delay in reporting of one day (green), with variations having higher or lower capacities or delays (see legend). Error bars represent SEM values over 5000 replicate simulations.

### Implementation of a saliva screening program

In the light of these results, we implemented saliva-based screening in Bangalore Life Science Cluster (BLiSC) campus of ca. 1400 adults. Participants donated a saliva sample once every seven days, and information was collected using a mobile phone application. Individuals were instructed on the sample collection protocol using a video played at the collection centre with no verbal instruction. Samples were tested on the same day, the maximum delay between sample collection and testing being ca. 8 hrs. Over six months, we tested ca. 20000 saliva f samples for SARS-CoV-2 using the protocols described here. An average of 160 samples were tested each day, with a maximum of 300 samples on a single day. Samples were collected between 10 am and 2 pm Monday to Friday, processed in about 6 hrs, with final results delivered soon after (the same evening) via mobile phones. Over these six months, we noted only ca. 10 instances where amplification of the internal control (RP) did not occur, presumably due to inhibitory factors in the collected saliva. Over that same period, nine saliva samples tested positive. Of these, eight individuals were completely asymptomatic, and one had very mild generic symptoms suggestive of an upper respiratory tract infection. Among these nine individuals, three tested positive on NP swabs collected on the same day, one tested negative, and the remaining 5 declined further testing and preferred to isolate as per public health guidelines.

## Discussion

Saliva is emerging as an effective alternative sample type for SARS-CoV-2 testing, with very high sensitivity and specificity [3, 9]. However, N/OPS-VTM remain the more effective sample type for routine diagnosis. Within this context, we examine the efficacy of the saliva for direct RT-PCR and their potential for large-scale testing. Our results suggest that saliva is an excellent alternative to conventional N/OPS-VTM with a reasonable concordance. The overall result agreement between N/OPS-VTM and saliva was 75.4%, slightly lower than reported in recent studies [10, 11]. This reduction in result agreement could be attributed to the longer storage duration of samples, timing of sampling and severity of the disease [12]. Since saliva samples were collected without any stabilization media, RNA stability might have been compromised during the storage and freeze-thaw, resulting in lower positivity [13, 14]. When recalculated for samples with less than two days of storage, concordance (83%) and sensitivity (74.5%) were closer to those reported in several recent studies [4].

The sensitivity of saliva was lower than that of N/OPS-VTM. This could be because most samples were follow-up samples of admitted positive patients collected after a week of hospitalisation. Many would have been symptomatic for more than two weeks; however, this information was unavailable. Recent studies have found a higher percentage of viral positivity in saliva when collected within ten days of COVID-19 diagnosis [15, 16]. Delayed sample collection could have been a reason for the low saliva sensitivity in our study. Another important finding of this study is that the Ct value comparison of three viral genes for samples with positive result agreement showed no significant variation. This suggests that these samples would have been taken in the initial phase of the infection. It also indicates that both positive N/OPS-VTM and saliva samples had a similar viral load, suggesting saliva is a valuable alternative sample type for SARS-CoV-2 detection [17].

Testing of asymptomatic individuals in a workforce is a proactive approach that can help identify and isolate sources of SARS-CoV-2 infection. In choosing the testing protocol, one confronts a trade-off between test sensitivity on the one hand, and testing capacity and testing delay on the other. For example, Rapid Antigen Tests produce immediate results with low sensitivity, while the gold-standard N/OPS RT-PCR test achieves high sensitivity but with limited capacity and delayed results. Furthermore, even a 100% sensitive test cannot prevent spread, since only a fraction of individuals are sampled each day, and individuals in early stages of infection may not test positive. We explored these trade-offs using a simulation of infection transmission. We found that increased testing capacity and decreased delay more than offset decreased test sensitivity, in preventing the emergence of large infection clusters. In particular, higher testing capacity enabled a more rapid cycle for testing an entire workforce, increasing the chance that an infection missed in one cycle was picked up in the next.

The saliva-based protocol described here meets the criteria highlighted above, for effective workplace screening. It is non-invasive, simple and self-collected without any PPE and can be used directly for RT-PCR [18]. The SalivaDirect protocol does not require RNA extraction, a significant bottleneck in the testing workflow [3]. These unique features of saliva screening significantly reduce testing costs and complexity when compared to N/OPS-VTM testing although we have not performed a formal cost analysis. Adopting saliva-based screening could yield higher testing capacity and shorter delays in result reporting. In case of resource constraints, saliva pooling can also be employed to further reduce overall turnaround time and test burden [19, 20]. Given these benefits, as well as the observed concordance between N/OPS-VTM and saliva, we piloted saliva-based screening to detect COVID-19 infections in the BLiSC academic campus. We found that this screening and surveillance effort was successful, enabling the campus to remain functional during the ongoing COVID-19 pandemic in India. Regular screening, which depends heavily on participant compliance, allows the resumption of normal workplace activity with enhanced safety with respect to COVID-19 infection. The approach described here can be scaled up for routine surveillance efforts, and implemented in schools, offices, academic institutions, and residential apartments.

## Conclusion

While there is substantial evidence for saliva as a source sample for SARS-CoV-2 detection globally, we present here a comprehensive study where we validate saliva as a screening tool for SARS-CoV-2 in India. Simulations aimed at detecting infection clusters guided the setup of a regular saliva-based screening on an academic campus, given estimates of concordance between saliva and nasopharyngeal/oropharyngeal swabs. Finally, we highlight the successful implementation of such a strategy on an academic campus. We hope this study can serve as an example and provide guidelines for the setup of a rapid and efficient approach to safe functioning of large establishments, including schools and industries, in the coming years as the world continues to live through a pandemic.

## Data Availability

All data produced in the present work are contained in the manuscript

## Acknowledgements

This work was primarily supported by NCBS/TIFR core fund, and we acknowledge the support of the Department of Atomic Energy (DAE), Government of India, under Project Identification No. RTI 4006; and philanthropic support from the Azim Premji Foundation. We acknowledge the CCAMP-InDx program for facilitating the saliva collection receptacle and transport medium from Kriyamed (https://www.kriyamed.com/) for campus screening. We are grateful to COVID-19 testing laboratory staff for helping with sample processing and RT-PCR testing; Mr. Prem Chandra Gautam, technical and research services teams at NCBS for their assistance in implementing campus saliva testing. We thank Prof. Apurva Sarin and Prof. Colin Jamora for facilitating the project. ABR is supported by the SPM research fellowship by CSIR-HRDG. UR is a Senior fellow DBT-Wellcome Trust India Alliance (IA/S/16/2/502714). SM is a JC Bose Fellow of the Department of Science and Technology, Government of India, and a Margadarshi Fellow of the DBT-Wellcome Trust India Alliance (IA/M/15/1/502018).

## CRediT author statement

**B.R. Ansil:** Conceptualization, Data curation, Analysis, Writing - Original Draft, Writing - Review and Editing. **Carolin Elizabeth George:** Conceptualization, Data curation, Analysis, Writing - Original Draft, Writing - Review and Editing. **Sindhulina Chandrasingh:** Conceptualization, Data curation, Writing - Original Draft, Writing - Review and Editing.

**Ashwin Viswanathan:** Conceptualization, Analysis, Writing - Original Draft, Writing - Review and Editing. **Mukund Thattai:** Conceptualization, Methodology, Analysis, Writing - Original Draft, Writing - Review and Editing. **Padinjat Raghu:** Conceptualization, Methodology, Writing - Original Draft, Writing - Review and Editing. **Santhosha Devadiga:** Investigation, Data curation. **Arun Geetha Harikumar:** Investigation, Data curation, Writing - Review and Editing. **Pulleri Kandi Harsha:** Investigation, Data curation. **Indu Nair:** Conceptualization, Data curation. **Uma Ramakrishnan:** Conceptualization, Writing - Original Draft, Writing - Review and Editing, Supervision. **Satyajit Mayor:** Conceptualization, Writing - Review and Editing and Supervision.

## Conflict of Interest

The authors declare that they have no conflict of interest.

## Notes

### Competing Interest Statement

The authors have declared no competing interest.

### Funding Statement

This study was primarily supported by NCBS/TIFR core fund (Department of Atomic Energy, Government of India, under Project Identification No. RTI 4006)

### Author Declarations

The study was approved by the Institutional Review Board of Bangalore Baptist Hospital (BBB/IRB/2020/010), Institutional Human Ethics Committee, National Centre for Biological Sciences (NCBS/IEC-22/01, NCBS/IEC-26/03), Institutional Biosafety Committee, National Centre for Biological Sciences (TFR: NCBS:34IBSC/UR1).

