## Supplementary Material 1 for "Validating saliva as a biological sample for cost-effective, rapid and routine screening for SARS-CoV-2"

### Materials and Methods

*Sample collection strategy*

Clinical samples were obtained from patients of Bangalore Baptist Hospital between December 2020 and May 2021. Assuming a sensitivity of 70% and a specificity of 85% at 50% prevalence, a minimum sample size of 162 was calculated with a precision of 10% at a 95% confidence interval [1, 2]. Patients admitted with COVID-19 were the most common source of positive samples and people who came for pre-travel testing predominantly contributed to the negative samples. There were 175 prospective patients in this study, 78 females and 97 males. Except for one patient (aged 16 years) all the patients were adults, ranging from 18-80 years in age. There were 124 symptomatic and 51 asymptomatic individuals in the study group. The purpose of the study was explained to the patients and written informed consent was taken before obtaining clinical samples. From each consented individual, a Nasopharyngeal swab or Oropharyngeal swab (N/OPS) and up to 5 ml of saliva were collected. N/OPS were collected in VTM (N/OPS-VTM), as per the standard testing protocol. The patient was asked to wash their mouth with water and then allow the saliva to pool in the mouth for about 1 to 2 minutes, after which they were asked to collect the sample in a sterile plastic container without any storage or stabilisation medium. Each paired sample was given a unique biorepository number and transported to the COVID-19 testing laboratory at Institute for Stem Cell Science and Regenerative Medicine (inStem), Bangalore Life Science Cluster (BLiSC) for RT-PCR testing once a day. Samples were stored in a 4°C to 8°C refrigerator till transport. Upon arrival, samples were stored in a 4°C refrigerator in the biosafety laboratory. Storage time before processing the samples varied from 0 to 15 days with a mean of 4 days. In case of storage beyond two days, the samples were moved to a -20°C freezer within the biosafety laboratory.

*Sample processing and RT-PCR testing for SARS-CoV-2*

N/OPS-VTM samples were processed following the Indian Council of Medical Research (ICMR) approved protocol. 200 µl of VTM solution containing the N/OPS was aliquoted into a 2.2 ml deep well plate containing 150 µl of lysis buffer (provided with the Beckman Coulter RNA extraction kit) for inactivation. RNA was extracted from these inactivated samples using a Magnetic bead-based automated Viral RNA Extraction protocol by Beckman Coulter Life Sciences. For saliva, 50 µl of the sample was aliquoted to a 1.5 ml centrifuge tube containing 6.5 µl of 20 mg/ml Proteinase K. Samples were vortexed thoroughly and then incubated for 5 minutes at 95°C. These samples were directly used (without RNA extraction) as the template for RT-PCR detection [3]. Samples were tested for RdRP, E and N genes of SARS-CoV-2 using NeoDx-CoviDx™ mPlex-4R SARS-CoV-2 RT-PCR Detection kits. The human RNase P gene was considered as the internal control. Test results were evaluated based on positive and negative control sets along with the test samples. As per the manufacture's criteria, only samples showing Ct (Cycle threshold) values below 40 were considered as positives. Based on the manufacturer's protocol, only samples in which at least one of the three SARS-CoV-2 genes and the RNase P were detected were considered as positives. RNase P undetected samples were considered inconclusive.

*Modelling infection spread on a network*

The model: Individuals are placed on an NxN square lattice with periodic boundary conditions, with transmission restricted to nearest neighbours. Time is updated in one-day increments. The infection course in a single individual is implemented using an extended SEIR (Susceptible / Exposed / Infected / Recovered) model, with the Exposed and Infected states split into kE and kI sub-states, respectively. Transmission from an Infected individual to any neighbouring Susceptible individual occurs with probability p per day, shifting the latter to the Exposed state. Transitions between successive Exposed and Infected sub-states and then to the Recovered state occur with probability q per day. C individuals are tested per day, moving row-wise along the lattice and covering the whole network every $N^2/C$ days. The test produces a positive result with probability s (equivalent to test sensitivity) on any Infected individual, and with probability zero on Susceptible, Exposed and Recovered individuals. Test results are reported after a delay of D days. Individuals who test positive are immediately isolated; their neighbours are then tested, and isolated after a subsequent D days if they test positive. We initialise the system with a single Exposed individual, and trace the dynamics of transmission until no individuals are Exposed or Infected. We repeat the simulation M times and track the fraction of times the number of infected individuals is equal to or exceeds L (defined as a large cluster). We use the following parameters: kE = 2, kI = 3; p = 0.5; q = 0.075; NxN = 1600; M = 5000; L = 25. Test sensitivity, daily testing capacity, and delay in reporting (s, C, D) are varied. With these parameters, the average duration of the Exposed and Infected states is 4 and 6 days, respectively; and the probability of a cluster of size 100 is approximately 1% in the absence of testing and isolation. This corresponds to a situation in which other NPI measures (masking, social distancing, etc.) are already in place, so infections do not spread across the whole network.
