## Supplementary Material 2 for "Validating saliva as a biological sample for cost-effective, rapid and routine screening for SARS-CoV-2"

**Supplementary Table 1:** Binomial Generalized linear model (GLM) results for the variables expected to be associated with result agreement between paired N/OPS and saliva samples. Only 'storage (3-15 days)' emerged as a significant variable for result agreement between paired samples. Storage of prospective samples for more than two days significantly reduces the result agreement.

| | **Estimate** | **Standard error** | **z value** | **Pr(>\|z\|)** |
|---|---|---|---|---|
| **(Intercept)** | 1.345922 | 1.035721 | 1.3 | 0.194 |
| **Sex (Male)** | -0.312578 | 0.477695 | -0.654 | 0.513 |
| **Age** | -0.004419 | 0.017493 | -0.253 | 0.801 |
| **Storage daybins 3-15** | -1.216768 | 0.542706 | -2.242 | **0.025*** |
| **Severity** | 0.265303 | 0.58228 | 0.456 | 0.649 |
